# Plasma concentration and safety of lopinavir/ritonavir in patients with Covid-19: a short communication

**DOI:** 10.1101/2020.05.18.20105650

**Authors:** Laurent Chouchana, Sana Boujaafar, Ines Gana, Laure-Hélène, Lucile Regard, Paul Legendre, Celia Azoulay, Etienne Canouï, Jeremie Zerbit, Nicolas Carlier, Benjamin Terrier, Solen Kernéis, Rui Batista, Jean-Marc Treluyer, Yi Zheng, Sihem Benaboud

**Affiliations:** Regional Pharmacovigilance Center, Pharmacology Department, Cochin Hospital, AP-HP.Centre – Université de Paris. Paris, France; Pharmacology Department, Cochin Hospital, AP-HP.Centre – Université de Paris. Paris, France; Pneumology Department, Cochin Hospital, AP-HP.Centre – Université de Paris. Paris, France; Internal Medicine Department, Cochin Hospital, AP-HP.Centre – Université de Paris. Paris, France; Antimicrobial Stewardship, Cochin Hospital, AP-HP.Centre – Université de Paris. Paris, France; Pharmacy Department, Cochin Hospital, AP-HP.Centre – Université de Paris. Paris, France

**Keywords:** Covid-19, SARS-CoV-2, plasma concentration, lopinavir, therapeutic drug monitoring, drug safety

## Abstract

**Background:** Lopinavir/ritonavir has been proposed as off-label treatment for Covid-19, although efficacy have not been proven. It has previously been shown that lopinavir plasma concentration is dramatically increased in inflammatory settings. As Covid-19 may be associated with major inflammation, assessing lopinavir plasma concentration and its safety in Covid-19 patients is essential.

**Methods:** Real-world Covid-19 experience based on a retrospective study.

**Results:** Of 31 patients treated by lopinavir/ritonavir for Covid-19, we observed very high lopinavir plasma concentrations, increased of 4.6-fold (IQR, 3.6-6.2) with regards to average plasma concentrations in HIV. All except one patient were above the upper limit of the concentration ranges of HIV treatment. About one over four to five patients prematurely stopped treatment mainly secondary an adverse drug reaction related to hepatic or gastrointestinal disorders.

**Conclusion:** Lopinavir plasma concentrations in patients with moderate to severe Covid-19 were higher than expected, associated with the occurrence of hepatic or gastrointestinal adverse drug reactions. However, owing that high plasma concentration may be required for *in vivo* antiviral activity against SARS-CoV-2 as suggested by previous studies, it appears that, in the absence of adverse drug reaction, lopinavir dosage should not be reduced. Cautious is necessary as off-label use can be associated with a new drug safety profile.

## INTRODUCTION

Since early December 2019, a pandemic infectious disease due to a novel coronavirus, namely severe acute respiratory syndrome coronavirus 2 (SARS-CoV-2), is spreading all over the world. To date, no specific therapeutic agent has proven its clinical efficacy against this outbreak. However, due to the urgent need for a treatment, several antiviral drugs are being used off-label, including lopinavir/ritonavir (LPV/r).^1^ LPV is a human immunodeficiency virus (HIV) protease inhibitor approved since many years. LPV is prescribed in HIV in association with another protease inhibitor, ritonavir (RTV), used as a potent P450 cytochrome (CYP) 3A4 inhibitor in order to dramatically increase LPV plasma exposure.^2^ LPV/r has been proposed in previous coronavirus outbreaks in 2003 and 2012 due to SARS-CoV-1 and to Middle East respiratory syndrome coronavirus (MERS-CoV), respectively.^3,4^ It has also proven its potency to inhibit SARS-CoV-2 replication in vitro.^5^ Based on these results, this drug has been considered to be potentially useful in patients with SARS-CoV-2. To date, LPV has been assessed in only one randomized clinical trial in Covid-19, showing no benefit beyond standard care in critical patients.^6^ No therapeutic drug monitoring was performed for LPV/r to assess an ideal drug exposure. This retrospective cohort aimed to assess LPV/r plasma concentration and its safety in SARS-CoV-2 infection.

## MATERIALS AND METHODS

Data were collected from routine care of Covid-19 patients from a single center setting (Cochin Hospital, Paris). According to our local protocol at the time of the study, patients were eligible to receive a seven to ten days LPV/r treatment as specific anti-Covid-19 therapy if they (i) had a pneumonitis evocating a SARS-CoV-2 infection at CT scan and (ii) were requiring oxygen (minimal flow rate 2-3 liters per minute). Patients with ARDS at admission and requiring intubation were not included. Covid-19 was considered as confirmed or suspected if they had a positive or negative PCR, respectively. LPV/r tablets were swallowed (not crushed) and administration time was reported by nurses. Therapeutic drug monitoring was performed in a routine care setting within the first three days of therapy. Plasma samples were considered as peak (4 +/-2 hours after drug intake) or trough (12 +/-3 hours after drug intake). Plasma concentrations of LPV and ritonavir were quantified with high-performance liquid chromatography tandem mass spectrometry (Xevo TQD, Waters^®^), using BEH C18 analytical column (1.7µm, 1.7µm, 50*2.1 mm, Waters, Saint-Quentin, France) and a mobile phase composed of 60% of water (0.05 % formic acid, v/v) and 40% of methanol (0.05% formic acid, v/v). LPV/r treatment safety was assessed in a routine care setting. Cases of suspected adverse drug reaction were spontaneously notified to the Pharmacovigilance regional center. After case-by-case assessment by a senior pharmacologist, cases were reported to the French pharmacovigilance system. The study has been performed in accordance with the declaration of Helsinki and received approval by the Cochin Hospital Institutional Review Board (number 2020-08019).

## RESULTS

We included 31 consecutive Covid-19 patients receiving LPV/r regimen between March 18^th^ and April 1^st^ 2020. The median age of patients was 63 (interquartile range [IQR], 51-78) years and 71% were men (Table 1). Pulmonary injury at CT scan was mostly moderate to extensive. All patients had a positive nasopharyngeal swab SARS-CoV-2 PCR, except four (12%) who had a doubtful or negative result, contrasting with a typical chest CT scan. Median C-reactive protein (CRP) and IL-6 levels at admission were 94.1 (45.4-176.0) mg/L and 60.4 (29.7-164.5) ng/mL. LPV/r 400/100 mg in tablets twice a day was started within the first 48 hours after hospitalization and median time to starting LPV/r after symptom onset was 8 (IQR, 7-10) days. No patient had a detected drug-drug interaction involving RTV and 13 (42%) patients did not received concomitant therapy. LPV/r treatment duration was 7 (IQR, 3-8) days. At the end of the LPV/r course, 17 patients were still hospitalized with oxygen dependency, eight patients were transferred to intensive care unit or died, five patients had recovered and clinical outcome was unknown for one patient that has been transferred to another hospital during treatment.

**Table 1.** Demographic and clinical characteristics of the patients.

| **Patient characteristics** |  |
| --- | --- |
| Age – year | 63 (51-78) |
| Sex – male:female | 22:9 |
| Comorbidities |  |
| Hypertension | 11 (35%) |
| Diabetes | 8 (26%) |
| Cardiovascular diseases (others) | 7 (23%) |
| Malignancy or immunosuppression | 6 (19%) |
| Chronic respiratory disease (including asthma) | 3 (10%) |
| Hepatitis or liver cirrhosis (Child-Pugh B or more) | 2 (6%) |
| Rheumatic disease | 2 (6%) |
| Chronic kidney failure | 1 (3%) |
| None | 6 (19%) |
| SARS-CoV-2 PCR |  |
| - Positive | 25 (80%) |
| - Doubtful* | 2 (6%) |
| - Negative* | 2 (6%) |
| - N/A | 2 (6%) |
| Extent of pneumonia at CT scan at admission |  |
| - Minor (less than 10%) | 1 (3%) |
| - Moderate (between 10-25%) | 9 (29%) |
| - Extensive (between 25-50%) | 12 (39%) |
| - Severe (more than 50%) | 7 (23%) |
| - N/A | 2 (6%) |
| Oxygen saturation at admission (without oxygen) – % | 92.5 (90-96) |
| C-reactive protein (CRP) at admission – mg/L | 94.1 (45.4-176.0) |
| Interleukin-6 (IL-6) at admission – ng/mL | 60.4 (29.7-164.5) |
| Time from symptom onset to starting lopinavir/ritonavir – days | 8 (7-10) |
| Type of Covid-19 related drug associated with lopinavir/ritonavir** |  |
| - None | 13 (42%) |
| - Cephalosporin or penicillin | 6 (19%) |
| - Cephalosporin and macrolide | 3 (10%) |
| - Macrolide | 3 (10%) |
| - Corticosteroids | 2 (6%) |
| - Corticosteroids and antibiotics | 2 (6%) |
| - Sarilumab (anti-IL-6) | 2 (6%) |
| Duration of lopinavir/ritonavir therapy – days | 7 (3-8) |
| Serum ALT level at the time of lopinavir assay*** |  |
| - < ULN | 13 (54%) |
| - 1-2 ULN | 7 (29%) |
| - 2-4 ULN | 3 (13%) |
| - 4-8 ULN | 1 (4%) |
| Reasons for therapy termination |  |
| - Scheduled end of treatment | 17 (52%) |
| - Adverse drug reaction | 7 (22%) |
| - Therapeutic limitation or deceased | 4 (13%) |
| - Poor efficacy | 3 (10%) |
| Types of adverse drug reaction accountable to lopinavir/ritonavir |  |
| - Cytolytic hepatitis | 3 |
| - Isolated hyperbilirubinemia | 1 |
| - Nausea and vomiting | 1 |
| - Diarrhea | 1 |
| - Agitation/anxiety | 1 |

Data are presented as median (IQR) or as no (%).
\* PCR resulting in doubtful or negative results have been repeated twice for each patient
\*\* penicillin: piperacillin/tazobactam; cephalosporin: cefotaxime; macrolide: azithromycin or
rovamycin
\*\*\* Serum alanine aminotransferase (ALT) level at the time of lopinavir assay +/- 1 day,
expressed in fold-changes above the upper limit of the normal (ULN) range. Data are provided
for the 24 patients included in the lopinavir therapeutic drug monitoring.

A total of 24 patients were analyzed for therapeutic drug monitoring; plasma assays were not available for three patients due to technical issues and four patients were excluded from analysis because of sampling out of peak or trough timings. Median time for peak and trough blood samples after tablet administration were 4,6 (IQR, 3,1-5,5) hours and 14,0 (IQR, 14,0-14,6) hours, respectively. LPV plasma concentrations ranges from 8,317 to 35,012 ng/mL. Median levels for peak (Cmax) and trough (Cmin) were 26,475 (IQR 11,952-33,868) ng/mL and 21,857 (IQR, 16,991-26,435) ng/mL, respectively (Figure 1A). All except one sampling were far above the upper limit of the concentration ranges observed in HIV. Extent of LPV plasma concentration was increased between 2 and 8-fold in the majority of the treated patients (Figure 1B). Magnitude of increase was not associated with CRP or IL-6 levels (Figure 1C, 1D). At the time of LPV assay ALT level was below 2 times the upper limit of normal (ULN) in 20 (83%) patients and between 2 and 8 ULN in 4 (17%) patients.

**Figure 1.**
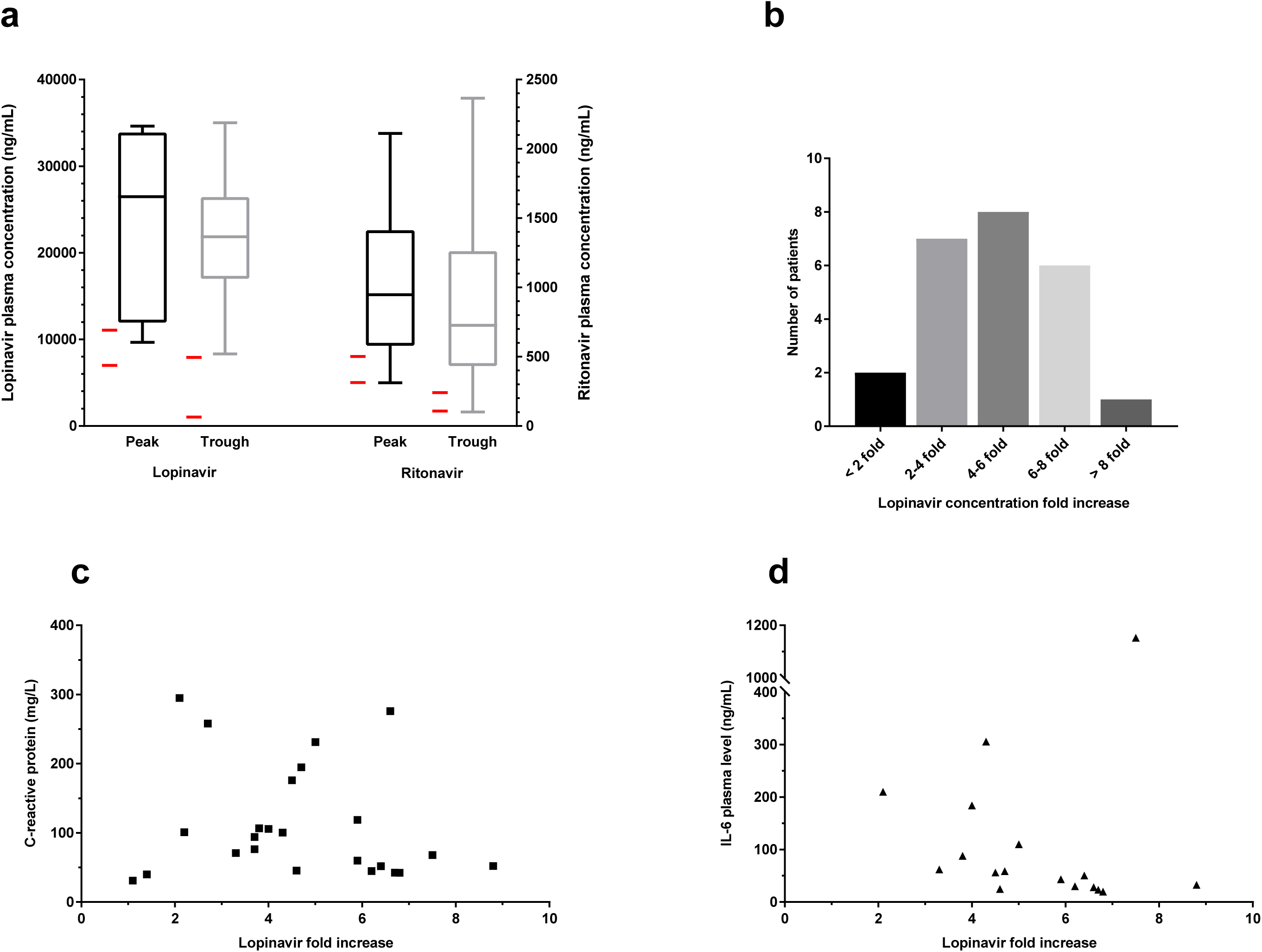
Lopinavir/ritonavir plasma concentrations and magnitude of lopinavir plasma concentration increase in Covid-19 patients compared to HIV patients. **a**. Plasma concentrations assayed at peak and trough in 6 and 18 patients with Covid-19, respectively. Boxes represent interquartile range and median, whiskers represent min and max values. Horizontal red line represent the peak and trough concentrations observed in HIV patients after 400 mg/100 mg lopinavir/ritonavir twice daily (i.e. for lopinavir at peak 7,000-11,000 ng/mL and at trough 1,000-8,000 ng/mL; for ritonavir at peak 300-500 ng/mL and at trough 100-250 ng/mL).^18,19^ **b**. Magnitude of lopinavir plasma concentration increase compared to average plasma concentration observed in HIV patients (i.e. 9,000 ng/mL at peak and 4,000 ng/mL at trough). **c** and **d**. C-reactive protein (r^2^=0.03) and IL-6 plasma levels (r^2^=0.02) according to the magnitude of lopinavir plasma concentration increase.

LPV/r was discontinued before the end of scheduled course for 14 (45%) patients (Table 1). Reasons for early ending LPV/r therapy were the occurrence of adverse drug reaction (n=7, 22%), therapeutic limitation or patient deceased (n=4, 13%) and poor efficacy (n=3, 10%). Suspected adverse drug reaction were assessed as probably related to LPV/r therapy. They consisted in four cases of liver injuries (three cases moderate cytolytic hepatitis between three and six times above the upper limit of normal range and one case of isolated hyperbilirubinemia), two cases of gastrointestinal disorders (nausea/vomiting and diarrhea) and one case of psychiatric disorders (agitation/anxiety). These adverse drug reactions were mild and all patients recovered after drug withdrawn.

## DISCUSSION

Our findings show that LPV plasma concentrations in Covid-19 patients were very higher than expected. Plasma concentrations were in median about 4.6-fold (IQR, 3.6-6.2) higher, and up to 8-fold higher, than therapeutic levels observed in HIV and far above the upper limit of the concentration ranges in all except one patient.

Major inflammation associated with elevated levels of blood IL-6 and NFκB upregulation have been largely reported in severe Covid-19 illness.^7^ Besides, it is long-standing known that inflammatory responses and infections impair drug metabolism capacity.^8^ In animals and humans, several studies or reports have shown the role of inflammation and IL-6 in CYP450 down-regulation through NF-κB activation, resulting in reducing drug metabolism.^9,10^ It has been previously reported that voriconazole or tacrolimus, highly CYP3A4 dependent drugs had their metabolism altered during inflammation. However, LPV is already associated with a highly efficient cytochrome inhibitor, ritonavir. It is unknown whether metabolism can be further blocked, and LPV concentrations increase through this mechanism. A prospective pharmacokinetics study in HIV patients also found that total LPV concentrations varied with inflammation and were correlated with circulating α□1□acid glycoprotein, a type 1 acute phase protein.^11^ Noteworthy, unbound LPV concentration were minimally altered during inflammation state suggesting a change in drug distribution rather than metabolism. In our analysis, we did not found a correlation between lopinavir levels and CRP levels measured at patient admission. Hence, it is likely that major inflammation in Covid-19 patients resulted in the dramatically increased LPV plasma concentrations found in our study. However, it is unknown if this results from a CYP3A4 reduced expression and another mechanism could be involved.

The main adverse drug reactions observed in this study were moderate hepatobiliary disorders that have been attributed to LPV/r therapy. In HIV, moderate-to-severe elevations in serum aminotransferase levels (>5 times the upper limit of normal) are found in 3% to 10% of patients.^12^ These elevations are usually asymptomatic and can resolve even with drug continuation. Drug causality could be hardly assessable in Covid-19 as up to 20% of the patients have increased transaminases.^13^ However, in our study, transaminases quickly decreased after stopping lopinavir, making drug causality probable. Furthermore, hepatobiliary disorders were mainly cytolytic with a moderate increase in serum transaminases. No patient had hepatic failure or dysfunction and it is not likely that these transient abnormal liver tests were associated with a decrease in hepatic metabolic ability. Overall, adverse drug reactions reported in this Covid-19 study are in line with LPV/r safety profile.^2^ Even though, due to the relative limited number of patients treated, conclusions are difficult to draw, physicians should be aware that drug off-label use can be associated with an altered drug safety profile.^14^ The drug is used in a setting not having been correctly assessed, that could led to an increase of adverse drug reactions and an unfavorable risk-to-benefits ratio.^15^

Antiviral activity assays on cultured Vero-E6 cells showed that the 50% effective concentration (EC_50_) of LPV on SARS-CoV and SARS-CoV-2 was 17.1 µM (i.e. 10,800 ng/mL) and 26.6 μM (i.e. 16,800 ng/mL), respectively.^5,16^ In our study, median trough plasma level was 20,153 (IQR 16,633-26,505) ng/mL barely at these concentrations or just above. The question of whether these plasma concentrations are effective to inhibit *in vivo* SARS-CoV-2 replication, especially in lungs, is unknown. Furthermore, Covid-19 associated vasculopathy or thrombosis could limit pulmonary diffusion. Altogether, this suggests that keeping high LPV plasma concentration appears essential considering a possible clinical efficacy of LPV in Covid-19.

Finally, as most HIV protease inhibitors, LPV has a very high protein binding.^17^ Therefore, in considering whether a dose is appropriate for SARS-CoV-2, it is critical to consider free drug concentrations. For standard HIV-1, free Cmin is 75 ng/mL that is substantially below EC_50_ reported for coronaviruses.^17^ Therefore, targeting SARS-CoV-2 antiviral activity in patients is challenging. It appears that, in the absence of adverse drug reaction, LPV/r dosage should not be reduced on the basis of therapeutic drug monitoring.

Our study has several limitations. First, due to the retrospective design using data from routine care, plasma assays were drawn at different times that prevent from an accurate pharmacokinetic estimation. Second, according to different practices in wards, Covid-19 severity at baseline was heterogeneous between patients. However, most importantly, in all assayed patients, LPV plasma concentrations regardless assays timing were unexpectedly high comparing HIV experience. Finally, unbound concentrations, which are the active ones, were not estimated.

## CONCLUSION

In conclusion, we found that LPV/r treatment in Covid-19 patients was associated with unexpected very high plasma concentrations. However, in the absence of adverse drug reaction, LPV dosage should not be reduced, owing that to high plasma concentration may be required for an *in vivo* antiviral activity as suggested by previous studies. We observed that about one over four to five patients withdrawn LPV/r therapy in relation with moderate adverse drug reactions. Cautious is needed in this context of drug off-label use, which can be associated with a new drug safety profile.

## Data Availability

Data are available upon request

## FOOTNOTES

Adverse drug reaction cases have been reported to the French pharmacovigilance system under the numbers PV20200174, PV20200224 and PV20200250-4.

## DECLARATIONS

### Ethics approval

The study has been performed in accordance with the declaration of Helsinki and received approval by the Cochin Hospital Institutional Review Board (number 2020-08019).

### Authors’ contributions

Study design: LC, SBo, IG, JMT, SBe

Data collection: LC, SBo, IG, LHP, LR, PL, CA, EC, JZ, NC, BT, SK, RB, JMT, YZ, SBe

Data analysis: LC, SBo; LHP, SBe Draft the manuscript: LC, SBo, SBe

Critically review the manuscript: LC, SBo, EC, NC, BT, SK

Approved the manuscript: LC, SBo, IG, LHP, LR, PL, CA, EC, JZ, NC, BT, SK, RB, JMT, YZ, SBe

